# EXTENDED ABSORPTION OF LIOTHYRONINE FROM POLY-ZINC-LIOTHYRONINE (PZL) IN HUMANS

**DOI:** 10.1101/2021.06.14.21258437

**Authors:** Alexandra M. Dumitrescu, Erin C. Hanlon, Marilyn Arosemena, Olga Duchon, Matthew Ettleson, Mihai Giurcanu, Antonio C. Bianco

**Affiliations:** Section of Adult and Pediatric Endocrinology and Metabolism, and Department of Public Health Sciences, University of Chicago, Chicago IL

**Keywords:** liothyronine, slow-release, metal coordination, hypothyroidism, thyroid, thyroxine, serum T3, serum TSH, thyroid hormone, PZL

## Abstract

**Background:** Liothyronine (LT3) has been increasingly used in combination with levothyroxine (LT4) in the treatment of hypothyroidism. A metal coordinated form of LT3, known as poly-zinc-liothyronine (PZL), avoided in rats the typical T3 peak seen after oral administration of LT3.

**Objectives:** To evaluate in healthy volunteers (i) the pharmacokinetics of PZL-derived T3 after a single dose, (ii) the pharmacodynamics of PZL-derived T3, (iii) monitoring for the adverse events; (iv) exploratory analysis of the sleep patterns after LT3, PZL or placebo administration.

**Methods:** 12 healthy volunteers 18 to 50 years of age were recruited for a Phase 1, double-blind, randomized, single-dose placebo-controlled, cross-over study to compare PZL against LT3 or placebo. Subjects were admitted three separate times to receive a randomly assigned capsule containing placebo, 50-mcg LT3, or 50-mcg-PZL, and were observed for 48h. A 2-week washout period separated each admission.

**Results:** LT3-derived serum T3 levels exhibited the expected profile, with a T_max_ at 2h and return to basal levels by 24-36h. PZL-derived serum T3 levels exhibited a ∼30% lower Cmax that was 1 h delayed and extended into a plateau that lasted up to 6h. This was followed by a lower but much longer plateau; by 24 hours serum T3 levels still exceeded ½ of C_max_. TSH levels were similarly reduced indistinguishably in both groups.

**Conclusion:** PZL possesses the necessary properties to achieve a much improved T3 pharmacokinetic. Drug product development of PZL should improve the delivery of T3 even further. PZL is on track to provide hypothyroid patients with stable levels of serum T3.

## Introduction

For decades, hypothyroidism was treated with desiccated extracts of porcine thyroid glands, which contain T4 and T3 (1)(2). With the development of TSH radioimmunoassay and the discovery that humans activate T4 to T3 (3), treatment with levothyroxine (LT4) became the standard of care (4)(5). Nonetheless, deiodination of T4 to T3 may not be sufficient to account for the normal thyroidal secretion of T3. First noticed around 1974 (6), subsequent studies revealed that LT4-treated patients maintain a ∼10% lower serum T3 levels (7,8).

Some LT4-treated patients complain of residual symptoms of hypothyroidism, such as poor quality of life, impaired cognition, difficulty with weight management, and fatigue, despite maintaining normal TSH levels (9-11). LT4-treated patients do weigh ∼10 pounds more (7), have a slower rate of energy expenditure (12), show slightly higher serum cholesterol levels (13), and are more likely to be on therapy to lower cholesterol levels (14). The extent of which these lower T3 levels contribute to the residual symptoms is unknown. In LT4-treated thyroidectomized rats, tissue euthyroidism only occurs after normalization of serum T3 (15). Such clear cut evidence is not available for LT4-treated patients. Nonetheless, the avialble data prompted professional task forces to recommend the development of a sustained-release T3 preparations (4,16).

Liothyronine (LT3) has been commercially available since 1956, but a lack of long-term safety data has limited its use (4). The pharmacokinetics (PK) of current LT3 products also raises potential concerns (17-19). A tablet of LT3 causes a T3 peak 2-3h after dosing, which sometimes is associated with palpitations, depending on the dose of LT3. Notwithstanding, little evidence exists of adverse reactions (AR) to appropriate doses of LT3 and a growing number of endocrinologists are prescribing LT4 plus LT3 to treat hypothyroidism. The analysis of ∼1,000 hypothyroid patients on combination therapy for up to 1 year did not reveal an increased frequency of AR when compared to LT4-treated patients (2). A retrospective analysis of 400 patients on LT3 for several years also did not reveal concerning trends (20). Thus, professional medical societies have become more accepting of combination therapy (4,16).

In pre-clinical studies we tested a new technology that employs metal coordination chemistry to produce a slow-release form of LT3. Coordinating T3 with metals under specific conditions yields a polymeric complex that adheres to the intestinal mucosal lining, creating a depot from which T3 is slowly released and absorbed over time. Studies in rats validated these concepts using oral administration of PolyZincLiothyronine (PZL) (21).

PZL is a prodrug that contains T3, which acts as a tridentate ligand binding to zinc via the participation of the amino acid group and the phenol group. When the phenol group is deprotonated, it acts as an additional zinc-binding site, thereby expanding coordination mode and favoring the formation of supramolecular structures. Extension of the intestinal transit time results from the inherent mucoadhesive properties of supramolecular metal coordinated (MC) complexes and the slow release of T3 from the MC complexes via ligand exchange (e.g. hydrolysis). The released T3 is absorbed into the bloodstream as T3 would normally (22). MC molecules adhere to the mucosa by mechanisms that include coordinate covalent bonding, hydrogen bonding, halogen bonding, metal-halogen bonding, and electrostatic interactions (23).

Here we report the first double-blinded placebo-controlled cross-over study of PZL in volunteers, that investigates the PK and pharmacodynamics (PD) of PZL-derived T3 in humans.

## Material and Methods

This clinical trial was conducted under the Food and Drug Administration (FDA) exploratory investigational new drug (IND) application (IND-137796), and approved by the University of Chicago IRB and Clinical Research Center (CRC) committees (IRB20-1341).

### Objectives

i. To evaluate the PK of PZL-derived T3 after a single dose of PZL by repetitive measurements of serum T3 levels.
ii. To evaluate PD of PZL-derived T3 through monitoring of serum TSH and free T4 levels, as well as heart rate and blood pressure.
iii. To evaluate the occurrence of AR or adverse events (AE) to a single dose of PZL through clinical interview and examination, metabolic panels, electrocardiogram.
iv. Exploratory analysis of the sleep patterns.

### Investigational Plan

This is a Phase 1, double-blind, randomized, single-dose placebo-controlled, cross-over study to test PZL in healthy volunteers.

#### Recruitment and screening

Healthy men and women volunteers 18 to 50 years of age, who self-report sleeping at least 7-hrs/night but no more than 9-hrs/night, between 22:00 and 08:00, were recruited from the community through advertisement. BMI <30 was an initial inclusion criterion, however as it became difficult to identify females not taking oral contraceptives, a female with BMI 33 was included to have enough female representation in the study.

Initial screening involved a telephone questionnaire. Exclusion criteria included the use of steroids or any medications known to affect thyroid hormone (TH) absorption or metabolism, or thyroxine-binding globulin, dietary utilization of kelp, soy, and/or biotin, pregnancy, lactation, use of oral contraceptives, previous diagnosis of sleep disorders (including obstructive sleep apnea (OSA)), prediabetes or diabetes, history of endocrine dysfunction or psychiatric, cardiovascular, or eating disorders, gastrointestinal disease that could affect the absorption of T3, drug or nicotine use, habitual alcohol use of >2 drinks per day, caffeine intake of >500 mg per day, history of bariatric surgery, weigh >100Kg, dietary restrictions, night shifts, or crossed any time zones in the month before the study.

Subjects who passed the initial screening visited the CRC, where they consented to the study. To qualify, individuals were determined to be in good health based on the medical history, physical examination. They underwent a 12-lead electrocardiogram (ECG), complete blood count (CBC), complete metabolic panel (CMP), and thyroid function tests (TFTs), i.e. serum TSH, FT4, total T3, and thyroid antibodies to thyroperoxidase and thyroglobulin. Women underwent a screening pregnancy test. Participants were eligible if ECG, CBC, CMP, and TFTs were normal.

#### Investigational product, dosage, and delivery system

Identical, off-white, size-0 capsules (Capsugel® Vcaps®, Lonza CHI) coated for duodenal delivery of contents were prepared by Catalent Inc (San Diego, CA). They contained current good manufacturing practice (CGMP) grade placebo (sterile excipient powder mixed with CGMP grade zinc chloride), PZL (56-mcg), or Na-T3 (53-mcg) prepared by Synthonics, Inc. (Blaksburg, VA); the amounts of PZL and Na-T3 were calculated to contain 50-mcg LT3, based on previous clinical experience with LT3 (17) and recommended dietary allowance (RDA) of zinc (10 mg/day). Capsules were labeled using 3 different alpha-numerical codes, and delivered to the Investigational Drug Service Pharmacy at the University of Chicago (kept at 4°C until use).

#### Admission to the CRC

Each volunteer was assigned (blindly and randomly) to a pre-defined sequence of treatment arms. In the morning of the trial (overnight fasting), subjects were admitted to the CRC, received their treatment, and were observed for 48h in the CRC. Subjects had timed blood draws and were monitored 3 times a day for AR through vital signs and physical exam (including neurological screening); an ECG was obtained previous to discharge. After discharge, these individuals returned to the CRC for a 7-day follow-up visit, for physical examination, ECG, CBC, CMP. After a 2-week washout period, volunteers were readmitted to one of the remaining treatment arms and the cycle was repeated. Thus, all individuals completed the three treatment arms, separated by 2-week washout periods.

#### Pharmacokinetics

An intravenous catheter was inserted into a forearm vein and left in place for serial blood sampling collected at specified times post-treatment and sent to the central laboratory for T3, FT4 and TSH levels using standard clinical bioanalytical methods. A total of 13 blood samples were drawn: -30 min, -15 min, 1h, 2h, 3h, 4h, 6h, 8h, 12h, 18h, 24h, 36h, and 46h.

#### Pharmacodynamics

Systolic and diastolic arterial blood pressure, as well as pulse, were measured at 30-minute intervals during the first inpatient day from the non-dominant arm using ambulatory monitoring equipment (Oscar II, SunTech Medical Instruments).

Sleep timing, duration, and fragmentation were assessed using wrist actigraphy monitors (Actiwatch Spectrum, Philips Respironics, Bend OR, USA) (32). Actiwatches were worn during the 2-day inpatient sessions and for the subsequent 4 outpatient days for a total of 6 days. Our primary measure of habitual sleep timing was at the midpoint of sleep, but we also calculated nocturnal sleep duration and indicators of sleep quality (sleep efficiency, sleep fragmentation). For screening, the Pittsburgh Sleep Quality Index (PSQI) was administered to assess sleep quality (24) and the Horne-Ostberg questionnaire to assess chronotype (25). Participants also completed a set of four validated computerized visual analog scales (0 to 10 cm) to assess hunger, appetite, mood, affect, sleepiness, and impulsivity throughout the in laboratory sessions, as we have previously reported (26) (27).

#### ARs and AEs

An independent safety monitor (Dr. Richard Abrams, Rush University Medical Center, Chicago IL) accessed all data and knew the treatment assignment of each volunteer. Based on clinical experience with the use of LT3 (28-31), he conferenced with Dr. Dumitrescu after each study arm was completed and, in all cases, recommended the continuation of the studies.

#### Statistical methods

For the sample size calculation, we used summary statistics (mean, SD, and inter-quartile range) for C_max_ and T_max_ from a previous study for LT3 (17). Based on pre-clinical data, we expect PZL-derived T3 to exhibit a decrease of 30% in C_max_ (mean = 242) and an increase in T_max_ of ∼6h (mean = 8.5). Assuming an SD of 100 for C_max_, we found that a sample of 12 individuals can detect a 30% decrease or higher in the mean C_max_ with a power of 90%. We further found that this sample of 12 individuals can detect an increase of 1h or more in the mean T_max_ with a power of 90%. Data analysis was conducted using statistical software R, and mixed-effects regression models were fit using the R package lme4 (33).

## Results

13 individuals were screened and 12 ultimately consented and were enrolled in the trial (Table 1). They were 31.6±9.9 years old; 4 were females. 11 individuals completed the trial as planned, given that one individual missed a study arm and was excluded.

**Table 1.** Demographics of all enrolled volunteers.

| ID# | Sex | Age |
| --- | --- | --- |
| A | F | 20-24 |
| B | M | 20-24 |
| C | M | 20-24 |
| D | M | 25-29 |
| E | M | 35-39 |
| F | F | 20-24 |
| G | M | 35-39 |
| H | M | 40-44 |
| I | F | 30-34 |
| J | F | 45-49 |
| K | M | 20-24 |
| L | M | 30-34 |

### T3 pharmacokinetics

#### Descriptive statistics and two-way ANOVA

In the LT3-arm, T3 serum levels increased from a baseline of ∼110 ng/dl to a C_max_ of ∼300 ng/dl between 2-3h (2.4±0.53) after dose delivery (Table 2; Fig. 1A). T3 levels decreased from ∼300 to 150 ng/dl by 12h, and then much less (dropping to 130 ng/dl) for the next 12h (Table 2; Fig. 1A). It took ∼8h for the serum T3 levels to decrease to ½ of C_max_ (Table 2; Fig. 1A). These two distinct phases of elimination are compatible with a two-compartment model for this experimental system (17)(19). T3 levels returned to baseline between 36-46h (Table 2). Two individuals exhibited atypical T3 profiles (C_max_ delayed by 6-24h) for no identifiable reasons and were excluded from further analyses.

**Table 2.**
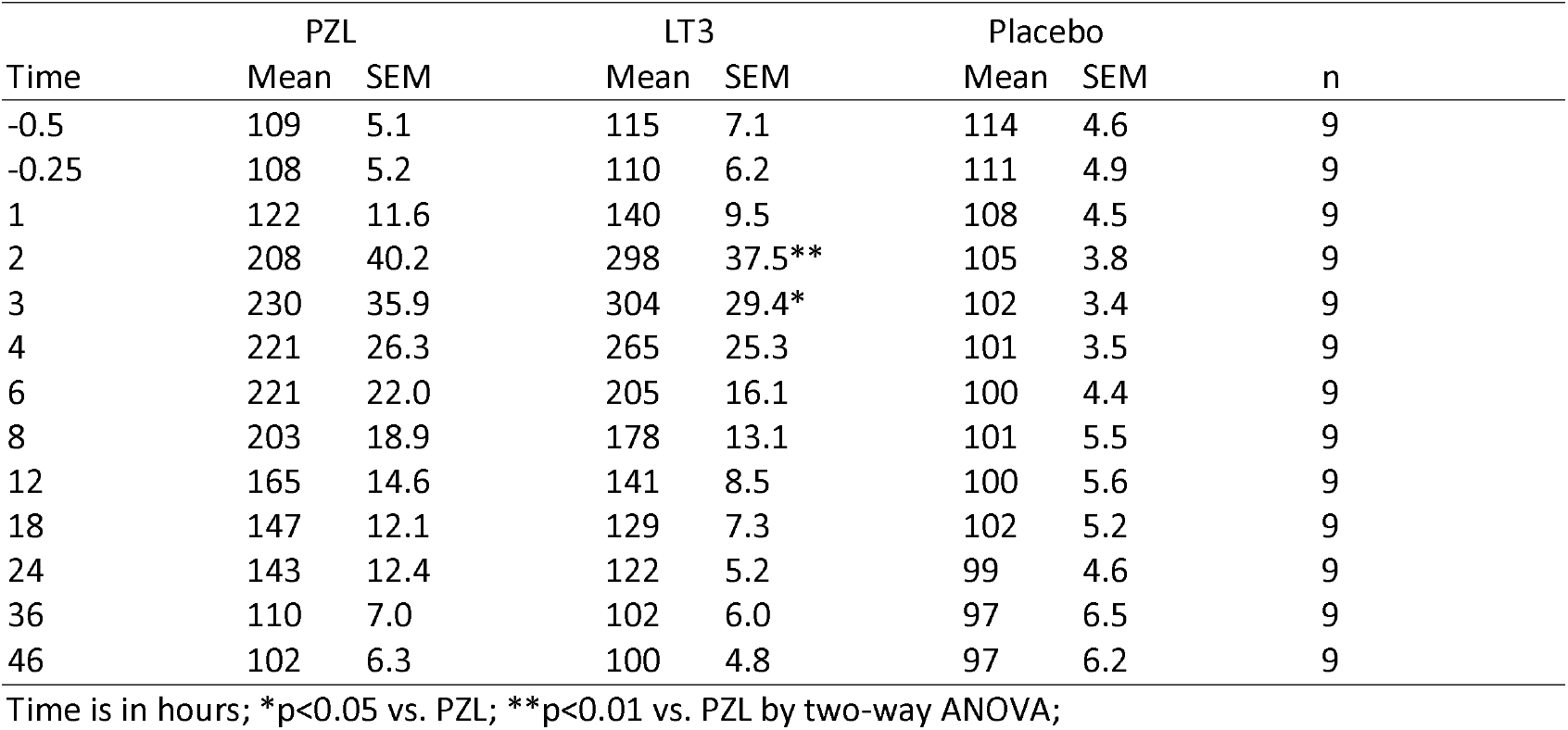
Serum T3 in volunteers during 48 h after taking a capsule of PZL, LT3, or placebo.

**Figure 1.**
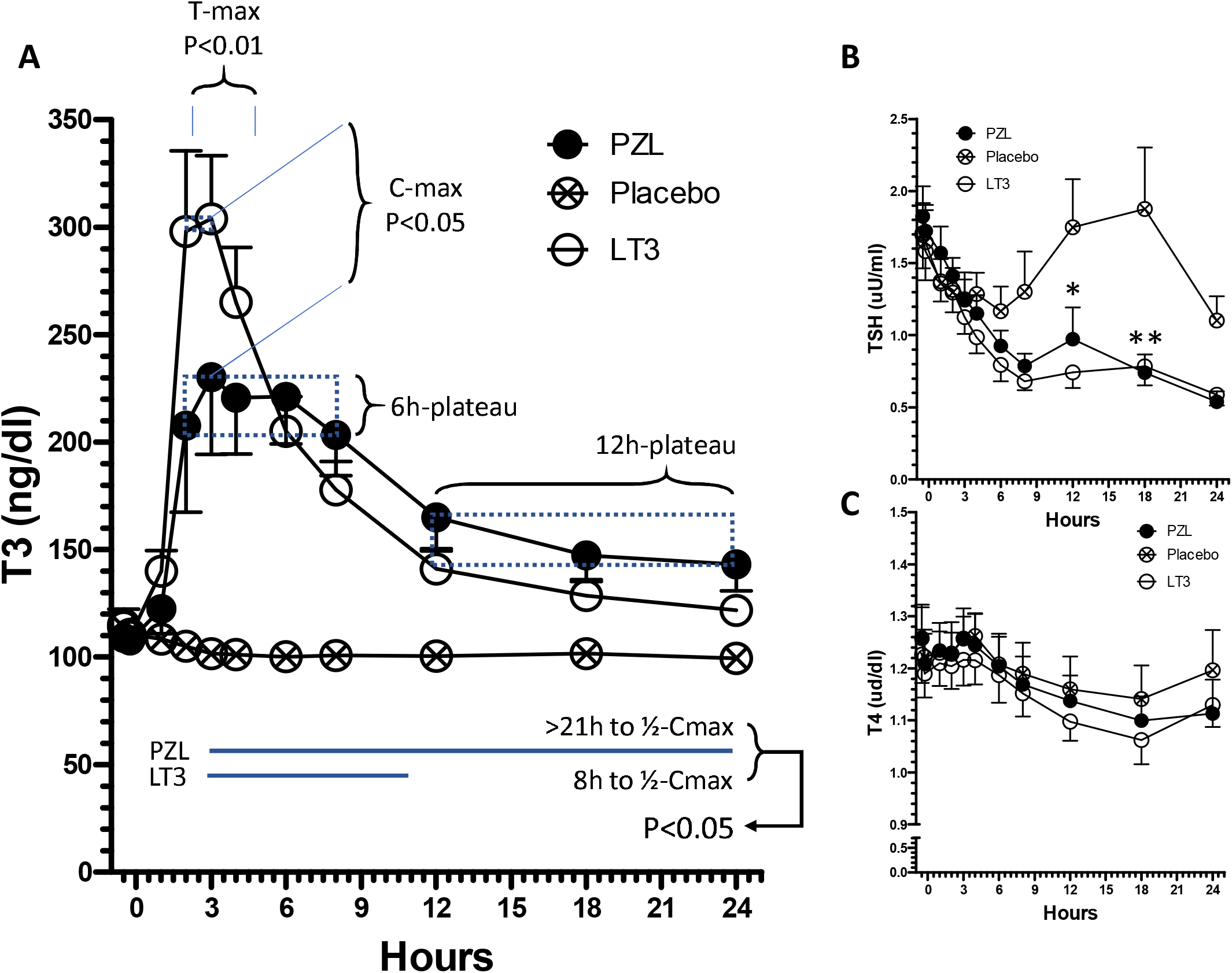
T3, T4, and TSH kinetics in volunteers after taking a capsule of PZL, LT3, or placebo. (A) Serum T3 values during the first 24 hours after treatments; T3 levels are stable between 2-3 hours after LT3 (p>0.05) but after PZL, T3 levels are stable for a longer time, between 2-8 hours (p>0.05); T3 concentrations at 2 and 3 hours are significantly different (two-way ANOVA followed by Bonferroni post-test); (B) Serum TSH values; *p<0.05 for both PZL and LT3 vs. place-bo; **p<0.001 for both PZL and LT3 vs. placebo; all statistics by two-way ANOVA followed by Bonferroni post-test; (C) Serum T4 levels.

In the PZL-arm, T3 serum levels increased from a baseline of ∼110 ng/dl to a C_max_ of ∼220 ng/dl between 2-8h (4.7±2.3) after dose delivery (Table 2; Fig. 1A). T_max_ was delayed in the PZL-arm by 1h (Table 2; Fig. 1A). T3 levels decreased slowly from ∼220 to 170 ng/dl up until 12h and then slowed down further, dropping to 150 ng/dl for the next 12h (Table 2; Fig. 1A). By 24h serum T3 levels still exceeded ½ of C_max_. T3 levels returned to baseline by 46h (Table 2).

In the placebo arm, T3 levels ranged from a baseline of ∼115 ng/dl to 100 ng/dl by end of the study (Table 2; Fig. 1A).

#### Mixed-effects models

We next fitted mixed-effects models for T3 levels under each time point considering patients as random effects to incorporate the repeated measures into the model. This model is suitable for individually randomized trials with longitudinal continuous outcomes. The analysis confirms that the T3 levels in the LT3-arm have a steeper slope and reach higher levels during the first 3h followed by a faster decrease when compared to the PZL-arm. In addition, starting at the 12h time-point, T3 levels were modeled at higher levels in the PZL-arm (Table 3).

**Table 3.**
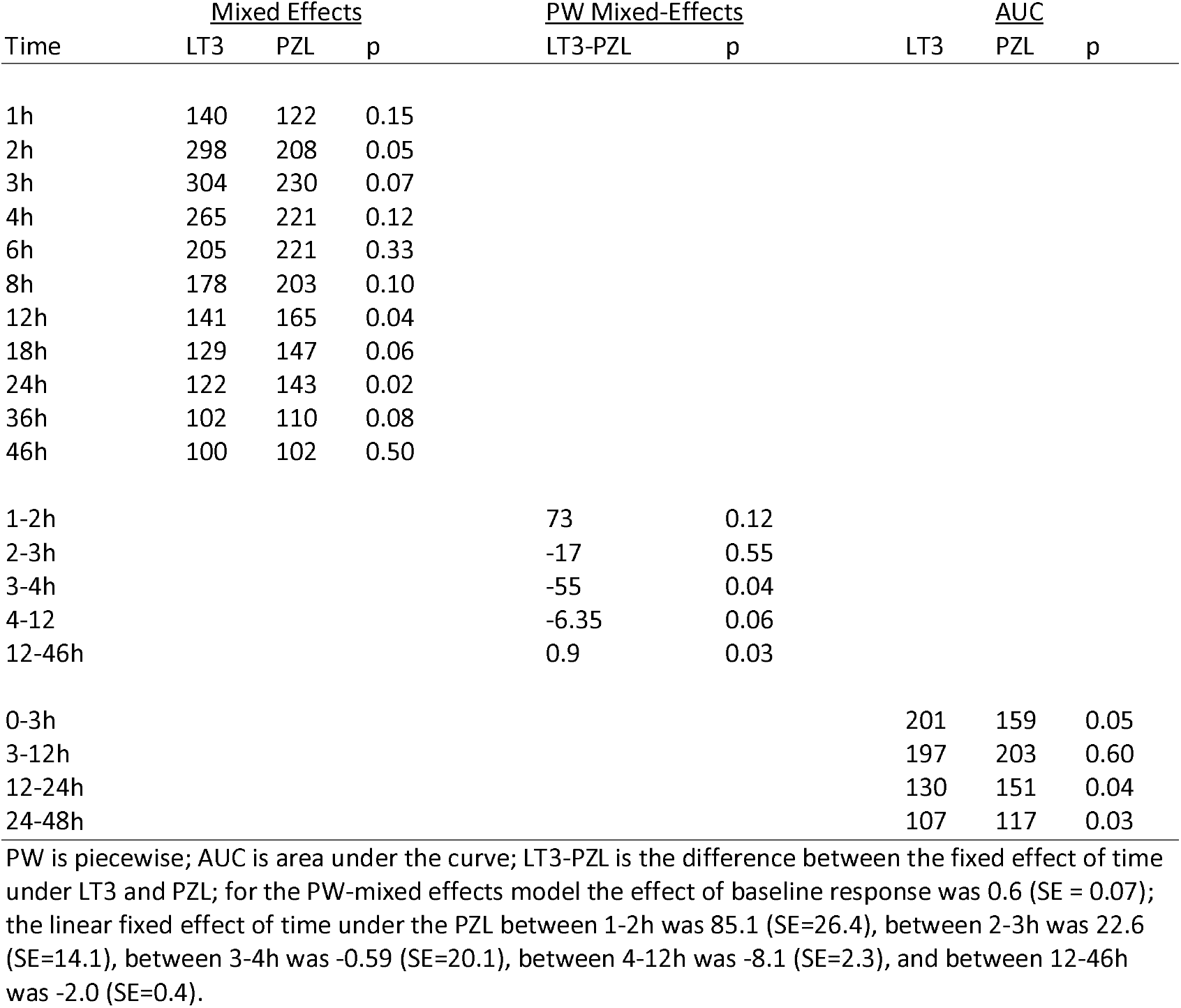
Mathematical modelling of LT3- and PZL-derived T3.

To study the integrated T3 levels, we calculated the area under each curve (AUC) for multiple time segments. It is clear that between 0-3h, the integrated T3 levels are higher in the LT3-arm, whereas from 12-48h they are higher in the PZL-arm (Table 3).

We next used the piecewise mixed-effects model with change points at 1h, 2h, 3h, 4h, and 12h, with average levels at -30 min and -15 min as a covariate and with random intercept and random slopes across the patients within each group. This model is useful when analyzing longitudinal data sets to model segmented change over time. It predicted a clear difference between the LT3 and PZL curves between 3-46h (Table 3).

### T3 pharmacodynamics

#### TSH and T4 levels

In the placebo-arm, serum TSH and T4 exhibited a reciprocal variation that reflects their normal circadian rhythmicity (Fig. 1B-C). Serum TSH dropped by ∼30% by 6h into the study only to return to baseline values by 18h, which was then followed by another ∼30% drop by 24h (Fig. 1B). At the same time, serum T4 levels varied much less, ∼10%, with reciprocal peaks and valleys (Fig. 1C). The elevation in serum T3 levels observed in the LT3-arm and PZL-arm provoked a similar reduction in serum TSH levels in both groups that reached ∼40% by 9h, and ∼70% by 24h, with disruption of the circadian rhythmicity (Fig. 1B).

#### Heart rate

There was a tendency for heart rate to increase during the day to a maximum around 4 PM, and then slowly return to baseline (Fig. 2), with no differences observed among treatment arms (Table 4).

**Figure 2.**
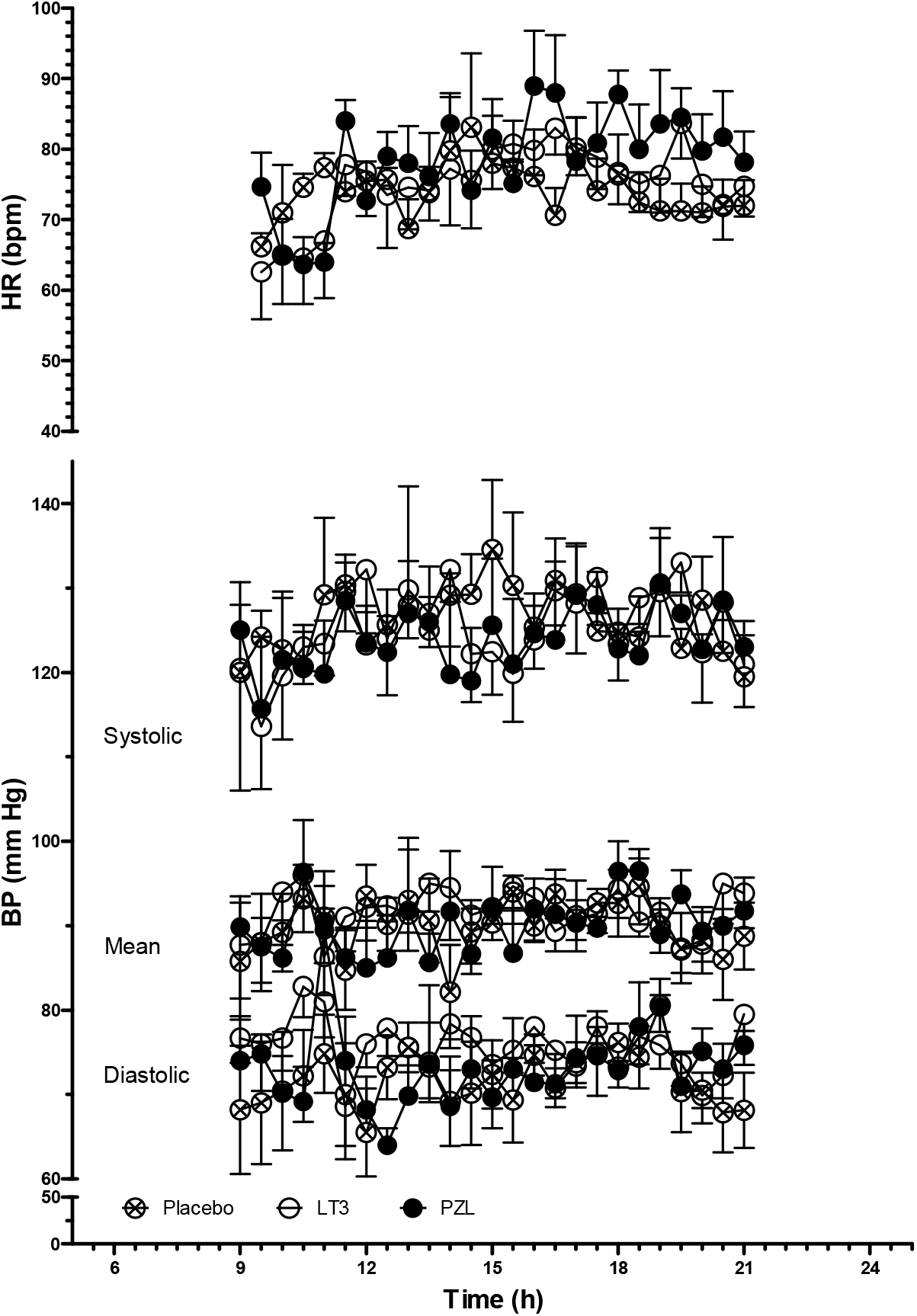
Heart rate, systolic, diastolic and mean blood pressures in volunteers after taking a capsule of PZL, LT3, or placebo.

**Table 4.** Heart rate in volunteers during the first 12 h after taking a capsule of PZL, LT3, or placebo.

|  | Heart Rate |  |  |
| --- | --- | --- | --- |
|  | PZ | T3 | PB |
| Number of values | 24 | 24 | 24 |
| Minimum | 64 | 63 | 64 |
| 25% Percentile | 75 | 74 | 75 |
| Median | 79 | 76 | 79 |
| 75% Percentile | 84 | 80 | 84 |
| Maximum | 89 | 84 | 89 |
| Mean | 79 | 75 | 79 |
| Std. Deviation | 7.0 | 5.6 | 7.0 |
| Std. Error | 1.4 | 1.1 | 1.4 |
| Lower 95% CI of mean | 76 | 73 | 76 |
| Upper 95% CI of mean | 81 | 78 | 81 |

BP is blood pressure; PZ is PZL, T3 is LT3, PB is placebo; the unit for heart rate is bpm; no statistically significant differences were observed by ANOVA

#### Blood Pressure

Systolic and diastolic blood pressures, as well as mean arterial pressure, remained stable throughout the day (Fig. 2), with no differences observed among treatment arms (Table 5).

**Table 5.** Systolic, diastolic, and mean blood pressure in volunteers during the first 12 h after taking a capsule of PZL, LT3, or placebo.

|  | Systolic BP |  |  | Diastolic BP |  |  | Mean AP |  |  |
| --- | --- | --- | --- | --- | --- | --- | --- | --- | --- |
|  | PZL | LT3 | PB | PZL | LT3 | PB | PZL | LT3 | PB |
| Number of values | 25 | 25 | 25 | 25 | 25 | 25 | 25 | 25 | 25 |
| Minimum | 115.7 | 113.6 | 119.5 | 64.00 | 65.50 | 68.60 | 85.00 | 82.14 | 86.33 |
| 25% Percentile | 121.3 | 122.3 | 123.1 | 70.00 | 69.27 | 73.47 | 86.71 | 88.13 | 89.89 |
| Median | 123.5 | 124.0 | 125.3 | 73.00 | 70.80 | 76.00 | 90.00 | 90.14 | 91.60 |
| 75% Percentile | 127.0 | 129.6 | 129.3 | 74.72 | 74.55 | 77.92 | 91.90 | 92.86 | 94.17 |
| Maximum | 130.6 | 133.0 | 134.6 | 89.50 | 80.57 | 82.80 | 96.50 | 94.63 | 96.00 |
| Mean | 123.9 | 125.6 | 126.2 | 73.18 | 71.83 | 75.77 | 90.09 | 90.01 | 91.66 |
| Std. Deviation | 3.659 | 4.919 | 3.886 | 4.786 | 3.346 | 3.180 | 3.372 | 3.163 | 2.746 |
| Std. Error | 0.732 | 0.984 | 0.777 | 0.957 | 0.670 | 0.636 | 0.675 | 0.633 | 0.549 |
| Lower 95% CI of mean | 122.4 | 123.5 | 124.6 | 71.20 | 70.45 | 74.46 | 88.70 | 88.70 | 90.52 |
| Upper 95% CI of mean | 125.4 | 127.6 | 127.8 | 75.15 | 73.21 | 77.08 | 91.49 | 91.31 | 92.79 |

BP is blood pressure; PB is placebo; the units for all BP values are mmHg; no statistically significant differences were observed by ANOVA.

### ARs or AEs

There were no deaths, no AR or serious AE. All AE were mild and resolved without complications; all were definitively not linked to LT3 or PZL treatment, as they were evenly distributed among arms (7 AE per arm). In the placebo-arm: anxiety, tiredness, faint episode, dizziness, back pain, knee pain, neck stiffness; in the LT3-arm: cold upper extremity, headache, blurry vision, nausea, nosebleed, low-grade fever, dizziness; and in the PZL-arm: dizziness, tiredness, low-grade fever, loose stools, sluggishness, headache (2x), sore throat. One woman had a delayed period during the trial with subsequent periods being on time, and another woman reported a heavier period in the month after the completion of the trial. No pregnancy occurred for any of the volunteers for the duration of the trial or during the 1 month afterwards.

All CBCs, CMPs, and ECGs obtained before, during, and 1 week after each admission arm were unremarkable (exceptions below). One volunteer had positive thyroid autoantibodies in presence of normal TFTs. One volunteer had slight elevation in total bilirubin of 1.2 mg/dL (0.1 – 1.0) at screening and ranged from 0.9 – 2.5 mg/dL during the trial, with the higher level of 2.5 obtained during the placebo-arm. Three women had low normal RBC at screening, 4.49×10^6^/uL, 4.82×10^6^/uL and 4.84×10^6^/uL (normal: 4.47-5.91×10^6^) and developed lower RBC during the trial, 3.98×10^6^/ulL 4.09×10^6^/uL and 4.38×10^6^/uL, respectively. A fourth woman and one man had low RBC at screening 4.23×10^6^/uL and 4.41×10^6^/uL, respectively, both reaching a lower level of 3.97×10^6^/uL during the trial. While the four women maintained a hemoglobin level in the normal range, the man with low RBC reached a lower hemoglobin level of 12.6 g/dL (normal range > 13.5 g/dL).

### Exploratory analysis of the sleep patterns

For all sleep outcomes, mean data was calculated for each participant from all days of collection, with a maximum of 6 contributing days within an arm. 10 participants completed actigraphy during the PZL-arm, of which 7 individuals had scorable data for all 6 days, 2 participants had 5 days, and 1 had 4 days contribute to the calculated means. 9 individuals completed actigraphy and had scorable data over all 6 days during the LT3-arm. For the Placebo-arm, 10 individuals completed actigraphy of which 9 had 6 six days of data and 1 participant had 3 days. No differences were observed in mean sleep outcomes including sleep onset, sleep offset, time in bed, sleep efficiency, and fragmentation among the groups (Table 6).

**Table 6.** Sleep parameters in volunteers during the first night after taking a capsule of PZL, LT3, or placebo.

|  | Units | PZL | LT3 | Placebo |
| --- | --- | --- | --- | --- |
| Bedtime | (min from midnight) | 11±72 | -2.9±57 | 12±62 |
| Wake time | (min from midnight) | 503±73 | 509±47 | 520±71 |
| Time in Bed | min | 492±37 | 512±49 | 509±40 |
| Assumed Sleep | min | 436±46 | 442±48 | 448±32 |
| Actual Sleep | min | 394±48 | 401±44 | 408±30 |
| Percent Sleep | % | 90±3.1 | 91±2.5 | 91±1.9 |
| WASO | min | 41±11 | 40±12 | 39±10 |
| Sleep Efficiency | % | 80±5.9 | 78±4.2 | 80±4.0 |
| Sleep fragmentation | %+% | 17±5.6 | 19±7.5 | 17±5.2 |

Numbers are the mean ± SD; for the time data, those are in minutes from midnight; bedtime is defined as the beginning of the rest interval, waketime is defined as the end of the rest interval, time in bed is defined as total minutes from bedtime to waketime, actual sleep is defined as the total number of epochs within the entire rest interval scored as sleep multiplied by the epoch length in minutes, percent sleep is the percentage of scored total sleep over the sleep interval multiplied by 100, WASO (wake after sleep onset) is defined as the total number of epochs scored as wake within the sleep interval multiplied by the epoch length in minutes, sleep efficiency is defined as the percentage of scored total sleep over the time in bed thus including time in bed before initial sleep onset and final awakening, and sleep fragmentation is defined as the sum of the percent mobile and percent immobile bouts as defined by actiware software for the entire rest interval.

## Discussion

Here we report the results of a phase-1 clinical trial in which a new polymeric metal-coordinated T3 molecule (PZL) was tested as a slow-release T3 formulation. In a double-blinded cross-over placebo-controlled trial, healthy volunteers were studied for 48h after they received capsules of placebo or equimolar amounts of T3 from LT3 or PZL.

Whereas the serum T3 profile observed after the LT3 dose was typical and consistent with previous studies, the serum T3 profile obtained after the PZL dose was substantially different. There was a 6h plateau in T3 levels around the C_max_ that was ∼30% lower when compared with LT3, which reflects the delayed but continued intestinal absorption of T3. At the same time, T3 levels in the circulation remained above ½-C_max_ for more than 24h, which was in contrast with the large swing in serum T3 levels observed after LT3 administration. This was confirmed with a mixed-effect and piecewise mixed-effects models, as well as sectional AUCs. The 6-hour plateau combined with the relative stability of T3 levels during the first 24h, set up an optimistic scenario for achieving more stable T3 levels during repeated administrations of PZL capsules at 24-hour intervals.

It is remarkable that despite the differences in PK, the reduction in serum TSH was similar in both LT3 and PZL-treatment arms. Serum T4 levels were only minimally affected in both cases. This is an indication that the prolonged and constrained elevation in serum T3 levels associated with PZL administration triggered similar cumulative PD effects, despite the absence of the marked peak of T3 levels in the circulation. Accordingly, the total AUC for serum T3 was similar for LT3 and PZL.

The volunteers were closely monitored before, during, and after each arm. Only minor AEs were observed, which were similar in frequency and intensity in all study arms. Even though the individuals were given 50 mcg LT3 (or the equivalent dose in PZL), and serum T3 levels increased substantially, no ARs were reported; no significant changes in heart rate or blood pressure were observed.

Professional medical associations have called for the development of slow-release formulations of LT3 (4) and for future trials to be performed with these new drugs (16). This is because the available studies on LT3 were not comprehensive, and in most cases were not designed to detect long-term ARs. Multiple strategies have been developed to answer those calls (34), including the oral administration of T3 sulfate (35,36).

The present study shows that a copolymer of T3 and zinc possesses the necessary properties to achieve a much improved T3 PK. This was a proof-of-principle study required by the FDA regulatory process. It is expected that even more consistent delivery of T3 will be achieved in the final steps of the drug product development, which includes refinement of the formulation with appropriate excipients and delivery method (i.e. coated tablets instead of enteric capsules). The successful development of PZL adds to the arsenal of new strategies and molecules that are being developed to provide stable replacement levels of T3 for patients that suffer from hypothyroidism.

## Data Availability

All relevant data are contained in the manuscript.

## Acknowledgments

We thank the director of the Clinical Research Center at the University of Chicago Medical Center, Dr. Arlene Chapman, clinical research nurses Kathy Reilly, Imani Wilson, Sally Pirowski, research coordinator Leeon Jones and bionutritional research manager Jennifer Kilkus for their dedicated help with the execution of the trial. Thanks are also given to the Kovler Diabetes Center executive director Peggy Hasenauer, clinical research coordinators Cristy Miles, Gail Gannon, Colleen Bender, Triniece Pearson and Rabia Ali, to Melanie Norstrom (Clinical Research Support Office, Department of Medicine), and to pharmacist Judy Pi (Investigational Drug Service Pharmacy Manager) for coordination of different aspects of the trial. We thank Dr. Richard Abrams from Rush University Medical Center for his valuable oversight as independent safety monitor. Research reported in this publication was supported by the National Institute Of Diabetes And Digestive And Kidney Diseases of the National Institutes of Health under Award Number R44DK116396. The content is solely the responsibility of the authors and does not necessarily represent the official views of the National Institutes of Health. AB is a consultant for Synhtonics, Allergan, Abbvie and BLA Technology.

## References

1. Chaker L, Bianco AC, Jonklaas J, Peeters RP. Hypothyroidism. Lancet. 2017.

2. Idrees T, Palmer S, Maciel RMB, Bianco AC. Liothyronine and Desiccated Thyroid Extract in the Treatment of Hypothyroidism. Thyroid : official journal of the American Thyroid Association. 2020;30(10):1399–1413.

3. McAninch EA, Bianco AC. The History and Future of Treatment of Hypothyroidism. Annals of internal medicine. 2016;164(1):50–56.

4. Jonklaas J, Bianco AC, Bauer AJ, Burman KD, Cappola AR, Celi FS, Cooper DS, Kim BW, Peeters RP, Rosenthal MS, Sawka AM. Guidelines for the treatment of hypothyroidism: prepared by the american thyroid association task force on thyroid hormone replacement. Thyroid : official journal of the American Thyroid Association. 2014;24(12):1670–1751.

5. Bianco AC, Salvatore D, Gereben B, Berry MJ, Larsen PR. Biochemistry, cellular and molecular biology, and physiological roles of the iodothyronine selenodeiodinases. Endocrine reviews. 2002;23(1):38–89.

6. Stock JM, Surks MI, Oppenheimer JH. Replacement dosage of L-thyroxine in hypothyroidism. A re-evaluation. The New England journal of medicine. 1974;290(10):529–533.

7. Peterson SJ, McAninch EA, Bianco AC. Is a Normal TSH Synonymous with “Euthyroidism” in Levothyroxine Monotherapy? The Journal of clinical endocrinology and metabolism. 2016:jc20162660.

8. Gullo D, Latina A, Frasca F, Le Moli R, Pellegriti G, Vigneri R. Levothyroxine monotherapy cannot guarantee euthyroidism in all athyreotic patients. PloS one. 2011;6(8):e22552.

9. Saravanan P, Chau WF, Roberts N, Vedhara K, Greenwood R, Dayan CM. Psychological well-being in patients on ‘adequate’ doses of l-thyroxine: results of a large, controlled community-based questionnaire study. Clinical endocrinology. 2002;57(5):577–585.

10. Wekking EM, Appelhof BC, Fliers E, Schene AH, Huyser J, Tijssen JG, Wiersinga WM. Cognitive functioning and well-being in euthyroid patients on thyroxine replacement therapy for primary hypothyroidism. Eur J Endocrinol. 2005;153(6):747–753.

11. Roberts ND. Psychological problems in thyroid disease.. British Thyroid Foundation Newsletter. Vol 181996.

12. Samuels MH, Kolobova I, Smeraglio A, Peters D, Purnell JQ, Schuff KG. Effects of Levothyroxine Replacement or Suppressive Therapy on Energy Expenditure and Body Composition. Thyroid : official journal of the American Thyroid Association. 2016;26(3):347–355.

13. McAninch EA, Rajan KB, Miller CH, Bianco AC. Systemic Thyroid Hormone Status During Levothyroxine Therapy In Hypothyroidism: A Systematic Review and Meta-Analysis. The Journal of clinical endocrinology and metabolism. 2018.

14. Idrees T, Prieto WH, Casula S, Ajith A, Ettelson M, Andreotti Narchi FA, Russo PST, Fernandes F, Johnson J, Mayampurath A, Maciel RMB, Bianco AC. USE OF STATINS AMONG PATIENTS TAKING LEVOTHYROXINE: AN OBSERVATIONAL DRUG UTILIZATION STUDY ACROSS SITES. J Endocr Soc. 2021;in press.

15. Werneck de Castro JP, Fonseca TL, Ueta CB, McAninch EA, Abdalla S, Wittmann G, Lechan RM, Gereben B, Bianco AC. Differences in hypothalamic type 2 deiodinase ubiquitination explain localized sensitivity to thyroxine. The Journal of clinical investigation. 2015;125(2):769–781.

16. Jonklaas J, Bianco AC, Cappola AR, Celi FS, Fliers E, Heuer H, McAninch EA, Moeller LC, Nygaard B, Sawka AM, Watt T, Dayan CM. Evidence-Based Use of Levothyroxine/Liothyronine Combinations in Treating Hypothyroidism: A Consensus Document. Thyroid : official journal of the American Thyroid Association. 2021;31(2):156–182.

17. Jonklaas J, Burman KD, Wang H, Latham KR. Single-dose T3 administration: kinetics and effects on biochemical and physiological parameters. Ther Drug Monit. 2015;37(1):110–118.

18. Saravanan P, Siddique H, Simmons DJ, Greenwood R, Dayan CM. Twenty-four hour hormone profiles of TSH, Free T3 and free T4 in hypothyroid patients on combined T3/T4 therapy. Exp Clin Endocrinol Diabetes. 2007;115(4):261–267.

19. Van Tassell B, Wohlford GFt, Linderman JD, Smith S, Yavuz S, Pucino F, Celi FS. Pharmacokinetics of L-Triiodothyronine in Patients Undergoing Thyroid Hormone Therapy Withdrawal. Thyroid : official journal of the American Thyroid Association. 2019;29(10):1371–1379.

20. Leese GP, Soto-Pedre E, Donnelly LA. Liothyronine use in a 17 year observational population-based study - the tears study. Clinical endocrinology. 2016;85(6):918–925.

21. Da Conceicao RR, Fernandes GW, Fonseca TL, Bocco B, Bianco AC. Metal Coordinated Poly-Zinc-Liothyronine Provides Stable Circulating Triiodothyronine Levels in Hypothyroid Rats. Thyroid : official journal of the American Thyroid Association. 2018;28(11):1425–1433.

22. Price JD, Piccariello T, Palmer S. Metal-coordinated pharmaceuticals. Drug development and delivery. 2014;14(1):34–39.

23. Smart JD. The basics and underlying mechanisms of mucoadhesion. Adv Drug Deliv Rev. 2005;57(11):1556–1568.

24. Knutson KL, Ryden AM, Mander BA, Van Cauter E. Role of sleep duration and quality in the risk and severity of type 2 diabetes mellitus. Arch Intern Med. 2006;166(16):1768–1774.

25. Roepke SE, Duffy JF. Differential impact of chronotype on weekday and weekend sleep timing and duration. Nature and science of sleep. 2010;2010(2):213–220.

26. Hanlon EC, Tasali E, Leproult R, Stuhr KL, Doncheck E, de Wit H, Hillard CJ, Van Cauter E. Sleep Restriction Enhances the Daily Rhythm of Circulating Levels of Endocannabinoid 2-Arachidonoylglycerol. Sleep. 2016;39(3):653–664.

27. Spiegel K, Leproult R, L’Hermite-Baleriaux M, Copinschi G, Penev PD, Van Cauter E. Leptin levels are dependent on sleep duration: relationships with sympathovagal balance, carbohydrate regulation, cortisol, and thyrotropin. J Clin Endocrinol Metab. 2004;89(11):5762–5771.

28. Celi FS, Zemskova M, Linderman JD, Smith S, Drinkard B, Sachdev V, Skarulis MC, Kozlosky M, Csako G, Costello R, Pucino F. Metabolic effects of liothyronine therapy in hypothyroidism: a randomized, double-blind, crossover trial of liothyronine versus levothyroxine. The Journal of clinical endocrinology and metabolism. 2011;96(11):3466–3474.

29. Celi FS, Zemskova M, Linderman JD, Babar NI, Skarulis MC, Csako G, Wesley R, Costello R, Penzak SR, Pucino F. The pharmacodynamic equivalence of levothyroxine and liothyronine: a randomized, double blind, cross-over study in thyroidectomized patients. Clinical endocrinology. 2010;72(5):709–715.

30. Tamai H, Fujino R, Shizume K, Kuma K, Suematsu H. [Changes in responsivity to TRH test and T3-suppression test after surgical treatment of hyperthyroidism (author’s transl)]. Nihon Naibunpi Gakkai Zasshi. 1975;51(12):985–996.

31. Stamp TC, Doar JW, Wynn V. Observations on some effects of L-triiodothyronine on carbohydrate and lipid metabolism in man. Journal of clinical pathology. 1969;22(2):132–135.

32. Jean-Louis G, von Gizycki H, Zizi F, Spielman A, Hauri P, Taub H. The actigraph data analysis software: I. A novel approach to scoring and interpreting sleep-wake activity. Percept Mot Skills. 1997;85(1):207–216.

33. Bates D, Maechler M, Bolker B, S. W. Fitting Linear Mixed-Effects Models Using lme4. Journal of Statistical Software. Journal of Statistical Software. 2015;67(1):1–48.

34. Idrees T, Price JD, Piccariello T, Bianco AC. Sustained Release T3 Therapy: Animal Models and Translational Applications. Front Endocrinol (Lausanne). 2019;10:544.

35. Santini F, Giannetti M, Ricco I, Querci G, Saponati G, Bokor D, Rivolta G, Bussi S, Braverman LE, Vitti P, Pinchera A. Steady State Serum T3 Concentrations for 48 Hours Following the Oral Administration of a Single Dose of 3,5,3’-Triiodothyronine Sulfate (T3S). Endocrine practice : official journal of the American College of Endocrinology and the American Association of Clinical Endocrinologists. 2014:1–25.

36. Santini F, Ceccarini G, Pelosini C, Giannetti M, Ricco I, Querci G, Grossi E, Saponati G, Vitti P. Treatment of Hypothyroid Patients With L-Thyroxine (L-T4) Plus Triiodothyronine Sulfate (T3S). A Phase II, Open-Label, Single Center, Parallel Groups Study on Therapeutic Efficacy and Tolerability. Front Endocrinol (Lausanne). 2019;10:826.

